# Estimates of COVID-19 deaths in Mainland China after abandoning zero COVID policy

**DOI:** 10.1101/2022.12.29.22284048

**Authors:** John P.A. Ioannidis, Francesco Zonta, Michael Levitt

## Abstract

**Background:** China witnessed a surge of Omicron infections after abandoning zero COVID strategies on December 7, 2022. The authorities report very sparse deaths based on very restricted criteria, but massive deaths are speculated.

**Methods:** We aimed to estimate the COVID-19 fatalities in Mainland China until summer 2023 using the experiences of Hong Kong and of South Korea in 2022 as prototypes. Both these locations experienced massive Omicron waves after having had very few SARS-CoV-2 infections during 2020-2021. We estimated age-stratified infection fatality rates (IFRs) in Hong Kong and South Korea during 2022 and extrapolated to the population age structure of Mainland China. We also accounted separately for deaths of residents in long-term care facilities in both Hong Kong and South Korea.

**Results:** IFR estimates in non-elderly strata were modestly higher in Hong Kong than South Korea and projected 987,455 and 619,549 maximal COVID-19 deaths, respectively, if the entire China population was infected. Expected COVID-19 deaths in Mainland China until summer 2023 ranged from 49,962 to 691,219 assuming 25-70% of the non-elderly population being infected and variable protection of elderly (from none to three-quarter reduction in fatalities). The main analysis (45% of non-elderly population infected and fatality impact among elderly reduced by half) estimated 152,886-249,094 COVID-19 deaths until summer 2023. Large uncertainties exist regarding potential changes in dominant variant, health system strain, and impact on non-COVID-19 deaths.

**Conclusions:** The most critical factor that can affect total COVID-19 fatalities in China is the extent to which the elderly can be protected.

## INTRODUCTION

China adhered to policies of “zero COVID” for almost three years during the COVID-19 pandemic with strictly enforced lockdowns and other aggressive restrictive measures. As of December 7, 2022, China decided to radically change course and adopt a strategy with few, more targeted measures. Restrictive measures were lifted abruptly and, as expected, wide circulation of Omicron variants ensued across the country. A major question is what might be the COVID-19 death toll in China under these circumstances. Projections, expectations and interpretations of accumulating data have started to abound. The Institute of Health Metrics and Evaluation (IHME) projected (1) 300,000 deaths until April 1, 2023 and 1 million deaths during 2023, with substantially different outcomes depending on use of masks and social distancing. In non-peer-reviewed announcements, some scientists, companies, media and social media commentators have projected extremely high death tolls with possibly up to several million fatalities (2-4). Concurrently, the official data of COVID-19 deaths from China have recorded very few deaths after removal of lockdown measures. Only 20 COVID-19 deaths were reported in the week of January 3-January 9, 2023. China is actually using a revised definition of COVID-19 deaths that requires the presence of pneumonia/respiratory failure due to SARS-CoV-2 infection and lack of comorbidities (3). This is a major change compared to previous recording of COVID-19 deaths, e.g. during the previous Shanghai outbreak practically all the recorded COVID-19 deaths occurred in people with comorbidities. This practice also deviates from COVID-19 recording practices in Western countries where both deaths ‘with’ and ‘by’ COVID-19 may be counted (5). Thus the official data from China and many speculations on COVID-19 deaths are becoming vastly divergent.

Here, we aim to offer estimates of the anticipated COVID-19 death toll in Mainland China from massive SARS-CoV-2 surges after the removal of lockdown measures. We employ comparative data from Hong Kong and South Korea. Hong Kong and South Korea witnessed large waves with Omicron variants starting in January 2022 after following until then policies that had maintained minimal infections in the population. We estimate the infection fatality rate (IFR) estimates across granular age strata in Hong Kong and South Korea and then extrapolate to Mainland China considering differences in the age structure of the population pyramids and in residents of long-term care facilities (LTCFs). We discuss also other potential differences that may affect comparisons between Mainland China in 2023 versus Hong Kong and Korea in 2022.

## METHODS

### Modeling of China on Hong Kong and South Korea Omicron waves

We aimed to estimate the COVID-19 death toll in Mainland China following the removal of lockdown measures on December 7, 2022 and until summer 2023. We used Hong Kong and South Korea as prototypes to model the expected deaths in Mainland China after adjustment of the age-structure in the respective population pyramids. Given that residents of LTCFs have particularly poor outcomes, we also took them separately into consideration.

Hong Kong witnessed a massive COVID-19 wave due to Omicron variants starting in January 2022. Seroprevalence data suggest that 45% of the community-dwelling population had been infected by mid-summer (6), 2.4 times the documented cases. We assumed the same ascertainment factor during 2022 with ascertainment factors per age strata informed by the seroprevalence study. Deaths of residents of LTCFs accounted for 54.5% of all COVID-19 deaths (7,8). As of December 27, 2022, after two new surges in the second half of 2022, the number of documented cases was 2,510,205 and the number of deaths was 11,349. BA.1 and BA.2 variants dominated until summer 2022 and BA.2 was gradually replaced by BA.5 and some BA.2.75, BQ1 and XBB variants later in the year (9).

South Korea similarly had the first Omicron deaths documented on January 3, 2022. Until the end of 2021, only 1.2% of the population had been documented to have been infected, while the proportion increased to 38.2% by July 31, 2022. By December 27, 2022 there were 31,882 deaths and 28,772,196 confirmed infections. South Korea uses aggressive testing and thus has very low rates of under-ascertainment, as corroborated also by limited seroprevalence data (10). Therefore, we assumed the true number of infections is 1.25-times the number of documented cases for each age stratum. BA.1 and BA.2 variants dominated until summer 2022 and BA.2 was gradually replaced by BA.5 and some BA.2.75 and XBB variants later in the year (9).

In China, the wave started shortly after removal of restrictive measures on December 7, 2022. For the first weeks, sequencing data suggest that the dominant variants are BA5.2 and BF.7 (11).

### Calculation of China COVID-19 deaths from age-stratified IFR estimates and population age structures

First, we projected the maximal COVID-19 fatalities in Mainland China, if the IFRs in different age groups (0-19, 20-29, 30-39, 40-49, 50-59, 60-69, 70-79, 80-, 90-) were the ones seen in Hong Kong or in South Korea during their respective Omicron waves and if the entire population of Mainland China were infected. Then, we considered different attack rates in the non-elderly (0-59 years old) population in China (ranging from 25% to 70%) until summer 2023, and different ability to protect especially the elderly above 60 year old (ranging from none up to reducing the fatality impact among them by three-quarters). The fatality impact in the elderly may be reduced either by preferentially shielding them from infection (12) or by further reducing the IFR e.g. by higher vaccination rates. In the main analysis scenario, we used 45% of the non-elderly population being infected by summer and fatality impact reduced by half in the elderly strata. Both Hong Kong and South Korea had approximately 45% of their population infected in the respective time frame in their Omicron waves (6,10).

For the highest age-stratum (above 80 years old), data on COVID-19 deaths are not reported separately for more granular substrata. However, the relative proportion of people 80-89 and above 90 years old can make a substantial difference, because IFR typically increases steeply with age (13,14). Therefore, we used a separate IFR for the population 80-89 years old (IFR_80-_/(4-(3xP_80-89_)) where IFR_80-_ is the IFR in the entire above 80 (>=80) age-stratum and P_80-89_ is the percentage of those 80-89 years old among those above 80 years old) and for those above 90 years old (IFR_80-_/(1-(0.75xP_80-89_))) in a way that the two estimates would differ 4-fold among them.

For Hong Kong, the majority of COVID-19 deaths during 2022 (54.5% of all COVID-19 deaths, 57% of COVID-19 in people above 60 years old) happened among the 76,091 residents of LTCFs (7,8) and their IFR was very high (probably because of very low vaccination rates in Hong Kong nursing homes) (15). Therefore, we generated separate IFR estimates for community-dwelling 60-69, 70-79, 80-89, 90-year old strata, and separately for residents of LTCFs. Assuming that 96% of deaths of residents of LTCFs are in people above 70 years old and the remaining 4% in people 60-69 years old, this corresponded to decreasing the community-dwelling IFR by 60% for people above 70 years old and by 25% for people 60-69 years old. For residents of LTCFs, we assumed that IFR was the same as the case fatality rate (9%) given the frequent, ubiquitous testing.

In South Korea, there are 24.9 beds in LTCFs per 1000 people over 65 years old (16). LTCFs were better protected with much higher vaccination coverage and accounted for a smaller share of deaths. Data have been published on infections and deaths per age stratum in residents of LTCFs for the 4 first months of 2022 in the province of Daegu-Gyeonbuk (17) and we used these as a proxy for the country, decreasing the community-dwelling IFR by 40% for people above 8 years old and by 20% for people 30-79 years old. For residents of LTCFs, we assumed that IFR was the same as the case fatality rate (606 deaths among 34,947 cases, i.e. 1.7341%) (17).

For China, it is estimated that there are 8.16 million beds for elderly people (18), but a large share of them are in day care facilities for overnight or short visits and the remaining are not fully occupied (19), therefore we assumed there are approximately 4 million residents of LTCFs.

### Population pyramids

Population pyramids for Mainland China, Hong Kong, and South Korea for 2022 were obtained from pyramidnet.net (20).

## RESULTS

### Calculation of stratified IFR for Hong Kong and South Korea

Tables 1 and 2 present the calculations of stratified IFR for Hong Kong and South Korea, respectively. Among community-dwelling non-elderly age strata (up to 59 years old), the IFR estimates were modestly lower in South Korea than in Hong Kong. IFRs were more similar among the community-dwelling elderly in the two locations. The IFR in residents of LTCFs were 5-times higher in Hong Kong than in South Korea.

**Table 1.** Calculations of stratified infection fatality rates in Hong Kong.

| Age | Population | Deaths | Cases | AsF | IFR | IFRc |
| --- | --- | --- | --- | --- | --- | --- |
| 0-19 | 1,277,620 | 18 | 325,661 | 3.3 | 0.000017 | 0.000017 |
| 20-29 | 798,498 | 19 | 288,699 | 2.4 | 0.000027 | 0.000027 |
| 30-39 | 1,104,277 | 31 | 418,923 | 2.1 | 0.000035 | 0.000035 |
| 40-49 | 1,141,564 | 79 | 414,715 | 2.3 | 0.000083 | 0.000083 |
| 50-59 | 1,169,327 | 329 | 397,424 | 2.3 | 0.000360 | 0.000360 |
| 60-69 | 1,122,469 | 969 | 352,545 | 2.5 | 0.001099 | 0.000825 |
| 70-79 | 608,372 | 1,869 | 170,189 | 2.5 | 0.004393 | 0.001757 |
| 80-89 | 288,088 | 7,979* | 112,863* | 2.5 | 0.016265 | 0.006506 |
| 90- | 94,091 |  |  | 2.5 | 0.065060 | 0.026024 |

Population as of 2022; deaths and COVID-19 cases during 2022 (fifth wave) as of December 27, 2022; 56 deaths and 29186 cases are not included as their assignment to age group was pending at the time of the analysis (see detailed data at Government of Hong Kong coronavirus site dedicated to the fifth wave statistics
https://www.coronavirus.gov.hk/eng/5th-wave-statistics.html).
AsF: ascertainment factor (ratio of total infections over documented cases) based on seroprevalence study in late July 2022 (reference 6).
IFR: infection fatality rate.
IFRc: infection fatality rate among community-dwelling population (excluding residents of long-term care facilities).
\*presented for all people aged 80 and over.

**Table 2.** Calculations of stratified infection fatality rates in South Korea.

| Age | Population | Deaths (2022) | Cases | IFR** | IFRc |
| --- | --- | --- | --- | --- | --- |
| 0-19 | 8,534,281 | 52 | 6,634,482 | 0.000006 | 0.000006 |
| 20-29 | 6,410,012 | 65 | 4,196,212 | 0.000012 | 0.000012 |
| 30-39 | 6,915,849 | 110 | 4,199,519 | 0.000021 | 0.000017 |
| 40-49 | 8,022,763 | 354 | 4,386,635 | 0.000065 | 0.000052 |
| 50-59 | 8,538,680 | 1029 | 3,731,215 | 0.000221 | 0.000177 |
| 60-69 | 7,044,982 | 2791 | 3,104,523 | 0.000719 | 0.000575 |
| 70-79 | 3,784,990 | 5742 | 1,572,122 | 0.002922 | 0.002338 |
| 80-89 | 1,788,653 | 16176* | 947,488* | 0.009631 | 0.005779 |
| 90- |  |  |  | 0.038524 | 0.023114 |

Population as of 2022; number of COVID-19 cases and deaths from the South Korea
government Central Disease Control Headquarters as of December 27, 2022
(https://ncov.kdca.go.kr/en/bdBoardList.do?brdId=16&brdGubun=161&dataGubun=&ncvCntSeq=&contSeq=&board_id=), subtracting the fatality impact per age group as of December 31, 2021 using historically archived data in Demography of COVID-19 database (https://dc-covid.site.ined.fr/en/data/korea/).
IFR: infection fatality rate.
IFRc: infection fatality rate among community-dwelling population (excluding residents of long-term care facilities).
\*presented for all people aged 80 and over.
\*\*assuming that cases in each age-stratum during the 5<sup>th</sup> wave is 1.25-times the number of cases recorded in the previous column.

### China projections of maximal deaths based on Hong and South Korea stratified IFR

As shown in Table 3, if the entire population of China were to be infected with IFR estimates similar to those obtained in Hong Kong and South Korea during 2022, the total COVID-19 fatalities would be 987,455 and 619,549, respectively. This included 119,630 and 59,946 fatalities, respectively, among non-elderly (0-59 years old) and 867,825 and 559,603 fatalities among elderly, respectively.

**Table 3.** Calculation of expected maximal deaths in China using Hong Kong or South Korea stratified IFR estimates.

| Age-group | China population | IFR (HK) | Maximal deaths<br>(per HK) | IFR (SK) | Maximal deaths<br>(per SK) |
| --- | --- | --- | --- | --- | --- |
| 0-19 | 335,323,469 | 0.000017 | 5,616 | 0.000006 | 2,103 |
| 20-29 | 175,108,854 | 0.000027 | 4,802 | 0.000012 | 2,170 |
| 30-39 | 233,767,164 | 0.000035 | 8,237 | 0.000017 | 3,919 |
| 40-49 | 202,957,964 | 0.000083 | 16,810 | 0.000052 | 10,482 |
| 50-59 | 233,838,693 | 0.00036 | 84,165 | 0.000177 | 41,273 |
| 60-69 (com) | 154,777,903 | 0.000825 | 127,626 | 0.000575 | 89,054 |
| 70-79 (com) | 81,730,213 | 0.001757 | 143,609 | 0.002338 | 191,047 |
| 80-89 (com) | 23,834,546 | 0.006506 | 155,068 | 0.005779 | 137,730 |
| 90- (com) | 3,132,598 | 0.026024 | 81,523 | 0.023114 | 72,408 |
| LTCFs | 4,000,000 | 0.09 | 360,000 | 0.017341 | 69,364 |
| Total |  |  | 987,455 |  | 619,549 |

China population as per 2022.
Com: community-dwelling population (excluding residents of LTCFs); HK: Hong Kong. SK: South Korea; IFR: infection fatality rate. LTCFs: long-term care facilities.
\*assuming 4 million residents of LTCFs distributed as 1 million 60-69, 1 million 70-79, 1.5 million 80-89, and 0.5 million above 90- years old.

### China projections of expected COVID-19 deaths until summer 2023

As shown in Table 4, the range of expected COVID-19 deaths in China until summer 2023 ranged from 49,962 to 691,219 under different scenarios. The most optimistic estimate uses the South Korean stratified IFR estimates and assumes that only 25% of the non-elderly population of China gets infected and the fatality impact among the elderly is reduced by three-quarters (e.g. by combination of factors such as better protection to avoid infection and/or better vaccination coverage). The most pessimistic estimate uses the Hong Kong stratified IFR estimates and assumes that 70% of the population is infected with no ability to offer any extra protection benefit to the elderly as compared with the non-elderly. In the main analysis where 45% of the non-elderly population is infected and fatality impact among the elderly is reduced by half, we estimated 152,886 to 249,094 COVID-19 deaths until summer 2023.

**Table 4.** Expected COVID-19 deaths in China until summer 2023.

| Percent<br>infected in<br>non-elderly | HK, S=1 | HK, S=0.5 | HK, S=0.25 | SK, S=1 | SK, S=0.5 | SK, S=0.25 |
| --- | --- | --- | --- | --- | --- | --- |
| 25 | 246,864 | 138,386 | 84,147 | 154,887 | 84,937 | 49,962 |
| 30 | 296,237 | 166,063 | 100,976 | 185,865 | 101,924 | 77,487 |
| 35 | 345,609 | 193,740 | 117,805 | 216,842 | 118,912 | 90,402 |
| 40 | 394,982 | 221,417 | 134,635 | 247,820 | 135,899 | 103,317 |
| 45 | 444,355 | 249,094 | 151,464 | 278,797 | 152,886 | 116,231 |
| 50 | 493,728 | 276,771 | 168,293 | 309,775 | 169,874 | 129,146 |
| 55 | 543,100 | 304,448 | 185,122 | 340,752 | 186,861 | 142,060 |
| 60 | 592,473 | 332,126 | 201,952 | 371,729 | 203,849 | 154,975 |
| 70 | 691,219 | 387,480 | 235,610 | 433,684 | 237,823 | 180,804 |

HK: based on Hong Kong-inferred infection fatality rates.
SK: based on South Korea-inferred infection fatality rates.
S=1, no precision shielding preferentially for the elderly (above 60 years old).
S=0.5, fatality impact reduced by half for the elderly.
S=0.25, fatality impact reduced by three-quarters for the elderly.

## DISCUSSION

An effort to estimate COVID-19 deaths in China until summer 2023 based on the respective experiences of Hong Kong and of South Korea during 2022 suggests a wide range of possibilities from ∼50,000 to ∼700,000, with values in the range of 150,000-250,000 for the main analysis. COVID-19 deaths in China are not a fixed, unavoidable calamity. How many people will die from SARS-CoV-2 depends primarily on the extent to which the elderly population can be protected and, to a much lesser degree, on the proportion of the non-elderly population that gets infected. It may also be further affected by whether the dominant variants during the epidemic wave have similar, lower, or higher fatality rates than the Omicron variants that dominated the 2022 outbreaks in Hong Kong and South Korea and by the functionality of the Chinese health system under strain.

The proportion of the population infected until summer 2023 in China may be higher or lower than the proportion of the population infected in the equivalent time span in 2022 in Hong Kong and South Korea (45%). The dominant variants in China as of end 2022 were BA 5.2 and BF.7 which may have become dominant because they are even more transmissible than the BA.1 and BA.2 variants that dominated in the first half of 2022 in Hong Kong and South Korea. This may lead to higher percentage of the population infected in China until summer 2023 and/or more explosive epidemic wave than may run its cycle faster. Conversely, differences in urbanicity and population density may point towards lower percentage of the population infected in China. Hong Kong is a highly densely populated metropolis and 81.4% of the population of South Korea is urban versus 64.7% in China. The extent to which restrictive measures, masks, and social distancing may be used in China (even without strict government mandates) adds to the uncertainty about the eventual attack rate.

The ability to selectively protect the elderly population is also uncertain. Evidence from seroprevalence studies from the first two COVID-19 waves suggests that some countries managed to offer substantial precision shielding to the elderly, but most countries had relatively similar infection rates among the elderly and non-elderly (21). With far greater sensitization about the extremely steep age risk gradient of IFR and with the past lessons of extremely high fatalities in LTCFs (22) elsewhere, including prominently in Hong Kong, one hopes that China avoids the past mistakes of other countries. Even though China has a smaller share of its population residing in LTCFs than Hong Kong and South Korea, a large share of avoidable deaths depends on whether these facilities can be effectively protected.

An even larger share of avoidable deaths pertains to community-dwelling elderly people. Vaccination coverage among the elderly as of end-2022 in China is better than the respective coverage of Hong Kong elderly and LTCF residents in early 2022. Only 25% of people over 80 years old had been vaccinated by January 2022 in Hong Kong (23), while in China as of December 2022 overall vaccination coverage is 92% and 40% of people over 80 years old have received 3 vaccine doses (24). The ability to offer vaccine boosters to all the very high-risk people, the availability of effective treatments and the pragmatic effectiveness of the vaccines used in China (25-27) will further shape the eventual death toll. CoronaVac may be modestly less effective than mRNA vaccines (26,27).

It is unclear whether the currently dominant BA.5.2 and BF.7 subvariants in China are less lethal than the BA.1 and BA.2 variants that dominated in Hong Kong and South Korea during the first half of 2022. Case fatality rates have only decreased slightly in South Korea between the first and second half of 2022 (when BA.5 became dominant). Conversely, case fatality rates have decreased substantially in Hong Kong during the second half of 2022 versus the first half. However, this may not be due to the dominance of BA.5, but due to better vaccination coverage and prior infections in a large segment of the population. The most likely scenario would be that for China as well, IFR may decrease during the course of 2023. However, long-term predictions are very uncertain, since the potential evolution towards new variants and the fatality thereof are unknown.

A comparison of stratified IFR estimates in Hong Kong, South Korea and China needs to consider also differences in the prevalence of risk factors for death after SARS-CoV-2 infection. China has a lower percentage of obesity in its population compared with Hong Kong and South Korea (28), but obesity has increased in recent years (29) and the difference may thus not be so prominent. Smoking rates (and thus respiratory disease) have been historically high in all three locations. The advantage of recent declines in smoking rates more prominently in Hong Kong probably do not translate yet to major decreases in severe respiratory disease. Transplants and other causes of medically-induced immunosuppression are less prevalent per population base in China. Overall, age-stratified IFR may be slightly or modestly lower in China than in Hong Kong and South Korea. If so, estimates of projected COVID-19 fatalities in China may be slightly or modestly overestimated.

Our main analysis estimates of COVID-19 deaths in China tend to be modestly lower than those proposed by IHME (1), but the range of possible outcomes with different scenarios amply overlaps. Our estimates are much lower than several non-peer-reviewed estimates that have been popular in media (2-4,30,31). For example, models by Wigram Capital Advisors (an investment firm), Airfinity (a data firm), and Economist (a magazine) anticipate 1 to 2.1 million COVID-19 deaths in China within just several months of abandoning zero COVID policies (30,31). These high estimates are widely speculative and depend on questionable modeling assumptions, typically from SEIR modeling. Many circulating estimates build on the work by Cai et al. that in spring 2022 estimated 1.6 million deaths after removal of ‘zero COVID’ policy in China (32). However, Cai et al. used the early data from Hong Kong (with higher IFR), did not differentiate age strata among the very elderly (e.g. 80-89, and over 90 years old) and did not adjust for different IFR in LTCFs. Moreover, their baseline scenarios assume not only higher IFRs but also higher percentage of the population infected. Even these overall pessimistic models, nevertheless, allow for much better outcomes, if there is widespread use of boosters and optimized treatment, with as low as 72,000 deaths according to Economist under an optimal scenario (30).

A major limitation for our analyses is the difficulty of prefiguring the health system performance during a massive COVID-19 outbreak in China. It is unknown if the stress will be proportionally higher in China than in Hong Kong or South Korea, although both Hong Kong and South Korea were stressed during the peak of the Omicron waves. High system stress may translate to less favorable outcomes both for COVID-19 deaths and for non-COVID-19 deaths (33,34). In this regard, both Hong Kong and South Korea have probably more functional health systems than China overall – but there is substantial variability across China. This could be reflected eventually on escalating excess death counts in particular in China. Excess deaths are a useful summary metric of the composite impact of the pandemic, its indirect effects and of the measures taken. However, reliable calculation of excess deaths requires accurate data on fatalities, ideally stratified per age group, over time. Gaps in death registration make these calculations very precarious. For example, in China a study conducted in 2018 found an average completeness of death registration of 72% with range from 2.4% to 100% across different counties (35). Even for South Korea, different modeling approaches yield different estimates of excess death. E.g. in 2020-2021, WHO estimates 6,288 excess deaths in South Korea (36), Economist estimates 7,558 excess deaths (37), while our calculations (using 2017-2019 as baseline) shows a death deficit (38). Economist projects 54,352 excess deaths as of November 14, 2022 (37), while our calculations based on age-stratified data (available until summer 2022) and using 2017-2019 as a baseline suggest a persistent death deficit until the summer of 2022. For China, uncertainties about excess deaths are likely to be even more prominent. E.g. Economist calculates over 700,000 excess deaths in China even before December 7, 2022 (37), i.e. with practically negligible COVID-19 deaths. Nevertheless, if COVID-19 deaths in China end up being in the range predicted by our main analysis, they would represent a very small percentage of overall mortality during the 2000-2023 period (<1% based on our main analysis), given that there are more than 10 million deaths annually in China.

Our analysis has several other limitations. First, seroprevalence surveys and estimation of ascertainment factors have uncertainty and potential biases (39,40). Second, data on LTCFs are available with different definitions and levels of accuracy across the three locations. Some imputations had to be made in the absence of detailed information. Therefore, our estimates need to be seen with extra caution. Third, in principle long-term projections for a dynamically evolving virus are highly precarious. Thus we discourage estimates extending beyond the first half of 2023 and major surprises during even the first half of 2023 are possible. Moreover, re-infections may have substantially lower IFR (41) and transmission dynamics may change in a heavily previously infected population. Fourth, relative estimates of effectiveness of various vaccines, e.g. the mRNA vaccines widely used in South Korea and the China-produced vaccines, are uncertain as they are based on observational data.

Finally, we used Hong Kong and South Korea as comparators for standardization, but other countries in the vicinity of China may also be informative. Japan and countries in Indochina already had substantial viral circulation and fatalities before 2022, in particular with Delta variants, so comparisons are less relevant. Taiwan adhered to zero COVID policy as well, but it abandoned this policy in late spring 2021 with its first massive Omicron wave starting 4 months later than Hong Kong and South Korea. It is unclear whether shifting the first massive exposure to the virus in late spring and summer rather than winter could make a difference. As of late 2022, it is estimated that 71% of the population of Taiwan has been infected (42) and 15,181 COVID-19 deaths have been reported as of December 28, 2022. We could not find detailed stratified data to perform analyses similar to Hong Kong and South Korea. However, the crude overall population IFR in Taiwan during 2022 (∼0.07%) is similar to South Korea’s.

Allowing for these caveats, our analysis provides a framework in which to attempt estimating COVID-19 deaths in China in the first half of 2023. Under most circumstances, the death toll may not be very high, but disparate estimates can emerge depending on the footprint of the epidemic wave on different population strata. The choice of China to report only a small subset of deaths compared to what has been reported by Western countries has drawn heavy criticism. It is unknown whether the intention is to reduce the potential for panic and the number of people who request healthcare attention, in particular in hospitals that may otherwise be overloaded with mildly symptomatic cases. Finally, while our analysis has an outlook of half a year, it is possible that in each city the duration of the epidemic wave will be much shorter, lasting only several weeks. The ability of the health system to absorb such acute waves without panic, the availability or not of resources and protection or lack thereof for the elderly and vulnerable may dictate the overall final outcomes of COVID-19 in China.

## Data Availability

All data are in the manuscript and tables. Archives of Hong Kong (COVID-19 fifth wave) and South Korea COVID-19 data can be found in https://www.coronavirus.gov.hk/eng/5th-wave-statistics.html and
https://ncov.kdca.go.kr/en/bdBoardList.do?brdId=16&brdGubun=161&dataGubun=&ncvContSeq=&contSeq=&board_id=

